# A high throughput splicing assay to investigate the effect of variants of unknown significance on exon inclusion

**DOI:** 10.1101/2022.11.30.22282952

**Authors:** Hilary A. Scott, Emily M. Place, Evelyn Harper, Sudeep Mehrotra, Broad CMG, Rachel Huckfeldt, Jason Comander, Eric A. Pierce, Kinga M. Bujakowska

**Author notes:** Correspondence: Kinga M. Bujakowska, Ph.D., Massachusetts Eye and Ear Infirmary, Harvard Medical School 243 Charles Street, Boston, MA 02114.

## Abstract

**Purpose:** Inconclusive interpretation of pathogenicity of variants is a common problem in Mendelian disease diagnostics. We hypothesized that some variants of unknown significance (VUS) may lead to aberrant pre-mRNA splicing. To address this we have developed a high throughput splicing assay (HTSA) than can be utilized to test the effects of 1000s of variants on exon recognition.

**Methods:** 2296 reference, control and variant sequences from 380 exons of 89 genes associated with inherited retinal degenerations (IRDs) were cloned as a pool into a split-GFP HTSA construct and expressed in landing pad RCA7 HEK293T cells. Exon inclusion led to disruption of GFP and exon skipping led to GFP reconstitution, enabling to separate GFP+ve and GFP-ve cells by fluorescence activated cell sorting. After deep sequencing-based quantification of studied sequences in each cell pool, exon inclusion index (EII) was determined, where EII = GFP-ve oligo count/total oligo count.

**Results:** HTSA showed high reproducibility when compared between different biological replicates (tetrachoric correlation coefficient r^2^ = 0.83). Reference exon sequences showed a high level of exon recognition (median EII = 0.88) which was significantly reduced by mutations to the essential splice sites (donor site variants: median EII=0.06; acceptor site variants: median EII=0.48). Of the 748 studied VUSs, 47 variants led to decreased exon inclusion (ΔEII ≤ −0.3) with 11 variants showing a strong effect (ΔEII ≤ −0,6). Using the HTSA data we were able to provide a likely genetic diagnosis to five IRD cases.

**Conclusion:** HTSA offers a robust method to study the effects of VUSs on exon recognition allowing to provide new diagnoses for patients with Mendelian disorders.

## INTRODUCTION

Despite substantial advances in next-generation sequencing technologies and their decreasing costs, genetic diagnoses are often hindered by our limited understanding of gene function, gene-disease relationship and uncertain pathogenicity of the uncovered variants^1–4^. For example, in the case of inherited retinal degenerations (IRDs), a Mendelian group of diseases affecting 2 million people worldwide^5^, the cause of disease in ∼30% of cases is unknown, despite substantial progress in genetic methodology and ∼280 known IRD genes^6–13^. The remaining missing diagnoses are in part due to the currently unknown IRD genes, however their contribution will be likely small as shown by the limited number of genes discovered within the last decade^14–34^. This suggests that the missing genetic causality largely lies in elusive pathogenic variation in the already known IRD genes, including structural and non-coding changes of the genome^1,35–45^. Since next generation sequencing (NGS) produces a vast quantity of variants, solutions to some of the unsolved cases may already be available in the existing data. We therefore need robust functional assays to test large numbers of variants uncovered in the remaining unsolved cases.

One of the ways that a variant can lead to disease is by causing aberrant pre-mRNA splicing, resulting in transcript degradation due to premature stop codons or deletion of a crucial protein domain^46^. These variants are not always changes of the canonical splice sites, but can be non-canonical splice site mutations in the proximity of the exon^47^, deep intronic variants^48–52^, synonymous^51,53^ or missense changes^54^. The pathogenicity of these variants is often difficult to predict even when using the recently developed machine learning algorithms designed for this purpose^55,56^. Due to the lack of certainty about the variants’ functional consequence, they are labeled as variants of unknown significance (VUS) or ignored altogether in the clinical diagnostic reports and research. Apart from having an effect on splicing, a VUS may affect transcription or transcript stability if they fall within the enhancer, promoter or the untranslated regions (UTRs)^57–59^. Also, despite the existence of a variety of pathogenicity prediction tools^60–64^, we lack sufficient data to interpret with confidence many of the missense variants, which can alter protein structure, enzymatic activity or interaction with other proteins or molecules. Our recent study of the genetics of IRDs showed that research “solutions” in ∼23% of cases are based on one or more VUSs^13,65^. All of these classes of VUSs require sophisticated variant class or protein specific assays^54,58,66,67^, which cannot be undertaken by a single laboratory. However, given the recent advances in the gene therapy field, an effort to study these variants is particularly important as it can finalize genetic diagnoses for patients, some of whom may be eligible for the emerging genetic therapies^68–74^.

In this study we wanted to address the contribution of VUSs to aberrant splicing^75–78^. To address this challenge, we developed a high throughput splicing assay (HTSA) to test the effect of hundreds of VUSs on aberrant splicing simultaneously. Our assay is an adaptation of a previously published methodology by Cheung and colleagues^54^ with an important alteration that allows for studying out-of-frame exons. We have tested our assay on 1131 variants in IRD genes and found likely genetic diagnoses for ten patients.

## MATERIAL AND METHODS

### Ethics declaration

The study was approved by the Institutional Review Board at the Massachusetts Eye and Ear (Human Studies Committee MEE, Mass General Brigham, USA) and adhered to the tenets of the Declaration of Helsinki. Informed consent was obtained from all individuals on whom genetic testing and further molecular evaluations were performed.

### Patient sequencing and annotation

Targeted panel sequencing was performed at MEE genomics core and exome sequencing was performed at the Center for Mendelian Genomics at the Broad Institute of MIT and Harvard using methodology described previously^79^. Sequencing data was aligned to human genome 38 and variants were called using Genome Analysis Toolkit (GATK) HaplotypeCaller package version 3^80^ (https://software.broadinstitute.org/gatk/), and annotated using VEP^81^ and VCFAnno^82^. CNV predictions were produced using gCNV^80^.

### Exon selection for splicing assay and oligo design

Internal exons in known IRD genes that were 200 base pairs (bp) or less were selected as possible candidates for the splicing assay, which allowed for inclusion of at least 40bp of 5’ intron (acceptor site), 30bp of the 3’ intron (donor site) and 15 bp flanking adapter sequences containing *AgeI* and *NheI* restriction sites. After selecting exons that were not longer than 200bp and did not include the *AgeI* and *NheI* restriction sites, we used an in-house software, Mendelian analysis toolkit (MATK)^79^ to prioritize rare variants in these exons (MAF <0.003) found in the unsolved IRD subjects. Additional rare variants in the *EYS* gene were taken from the gnomAD database^83^. In total 1131 variants in 380 IRD exons were selected (Tables S1 and S2). A pool of ssDNA oligos synthesized on silicone array-based method was purchased from a commercial vendor (TWIST Biosciences, USA).

### High throughput splicing assay (HTSA) design

We have adapted a published approach from Cheung and colleagues^54^, who kindly shared with us the *SMN1* intron backbone plasmid with split GFP-T2A-mCherry, the *Bxb1* integrase plasmid and the HEK2937-RCA7 cell line, that provide a chromosomal landing pad site which allows stable expression of splicing reporter library at the adeno-associated virus integration site 1 (AAVS1) safe harbor locus^54,84^. In our design, we altered the *SMN1* split GFP-T2A-mCherry plasmid, where we decoupled the mCherry and GFP expression to be able to study out-of-frame exons. For the uniform expression of both fluorophores, we introduced EF1alpha promoters in front of the split GFP and mCherry genes. We will refer to this altered vector as the HTSA vector. The vector also contains attB sites for integration into the *AAVS1* locus of the HEK293T-RCA7 landing pad cell line, as well as a promoter-less puromycin resistance cassette. HEK293T-RCA7 contain an attP site and the promoter that allows for the site-specific recombination of the HTSA insert and coupling of the promoter and the puromycin resistance gene.

### HTSA library design and cloning

We designed two oligo libraries: 1) control library containing 380 study exons with reference sequences, acceptor and donor site changes (1140 oligos in total); and 2) variant library containing 380 study exons with reference sequences, acceptor site changes, donor site changes and study variants (2296 sequences in total). Sequences that included the *AgeI* and *NheI* restriction sites were excluded. The library was amplified with Fwd-AgeI primer: 5’-ACGGCCAGTACCGGT-3’ and NheI-Rev primer 5’-GCTATGACCGCTAGC-3’ with a high fidelity polymerase (KAPA HiFi HotStart ReadyMix, Roche) for a maximum of 20 cycles and cloned to the HTSA vector by the *AgeI* and *NheI* restriction cloning.

### Cell culture and FACS

After NGS confirmation of the desired diversity (Novaseq 6000, Illumina), the libraries were co-transfected with a vector containing *Bxb1* integrase into the HEK293T-RCA7 landing pad cell line which allowed for the integration of the split GFP exons and mCherry reporter at the AAVS1 locus^54^. Since the integrated constructs enable expression of puromycin resistance gene in the landing pad cell line, the cells were selected with puromycin and maintained in culture and passaged for 20 days to ensure elimination of non-integrated plasmids. Thus the fluorescence signal observed in a single cell should come only from the integrated construct. After 20 days in culture, the cells were fluorescence activated cell sorted (FACS) (Sony MA900 Multi-Application Cell Sorter, Broad Institute Cell Sorting Facility) into GFP positive and GFP negative bins. All collected cells were mCherry positive. To ensure library diversity, at least 1000 cells per oligo in the library were sorted.

### Genomic DNA extraction and deep sequencing

gDNA extraction was performed with a commercial kit (Qiagen All-Prep Kit) and the integrated fragments were PCR amplified with HTSA-NGS-F primer 5’-CTATATATAGCTATCTATGTCTACCGGT-3’ and HTSA-NGS-R primer 5’-GGCTGGAACTCTTGCGCTAG-3’ with a high-fidelity polymerase (Primestar GXL Polymerase, Takara) for a maximum of 20 cycles. The amplicons from each fluorescent bin were multiplexed and deep sequenced with a targeted depth of 500 reads per oligo (250×250 pair-end reads, Novaseq 6000, Illumina).

### Data analysis and statistics

Test exon sequence reads in each fluorescence bin were determined by NGS and matched to the oligo sequences in the corresponding library, counted and normalized by coverage (reads per million in each pool) and the FACS fraction of the fluorescence bin. Average number of reads per sorted cell was established and sequences with coverage representing less than 50 cells (control library) or 150 cells (full library) across both pools were excluded. Exon inclusion index (EII) was calculated for each sequence, where EII = GFP-ve read count/(GFP negative + GFP positive read counts) (Supplemental Figure 1). The impact of the variant was calculated by the difference of exon inclusion of the variant and the reference sequence: ΔEII = EII_control_ - EII_reference_. The experiments were performed in duplicate, and an average of ΔEII values were calculated. Reproducibility of the assay was calculated from two biological replicates using the EII values for each sequence and simple linear regression analysis.

All figures were prepared with Graphpad Prism 8 and Biorender.

## RESULTS

### Optimization of the high throughput splicing assay (HTSA)

As the starting point of the assay design we used constructs developed by Cheung and colleagues for the multiplexed functional assay of splicing using Sort-seq (MFASS)^54^. In the original MFASS construct a GFP gene is split by 1000bp-long sequence derived from introns 7 and 8 of the *SNM1* transcript (NM_000344), into which test sequences can be cloned in a multiplexed fashion using the *AgeI* and *NheI* restriction sites (Figure 1A). In the MFASS design the split GFP and the control fluorophore mCherry were separated by a self-cleavage 2A peptide, which allowed for the study of in-frame exons only, which was an important limitation of the assay. To increase the number of possible exons to characterize, we have changed the construct by separating expression of the two fluorophores, which are now both under the ubiquitous *EF1*alpha promoter, allowing for testing out-of-frame exons and cryptic exons without compromising mCherry expression (Figure 1A). For every mini-gene transcript there are two outcomes: 1) an inserted test sequence is recognized as an exon, leading to GFP disruption and no green fluorescence; 2) an inserted sequence is not recognized as an exon, leading to full reconstitution of GFP and presence of green fluorescence (Figure 1B). The control mCherry fluorescence is present regardless of the splicing outcome.

**Figure 1.**
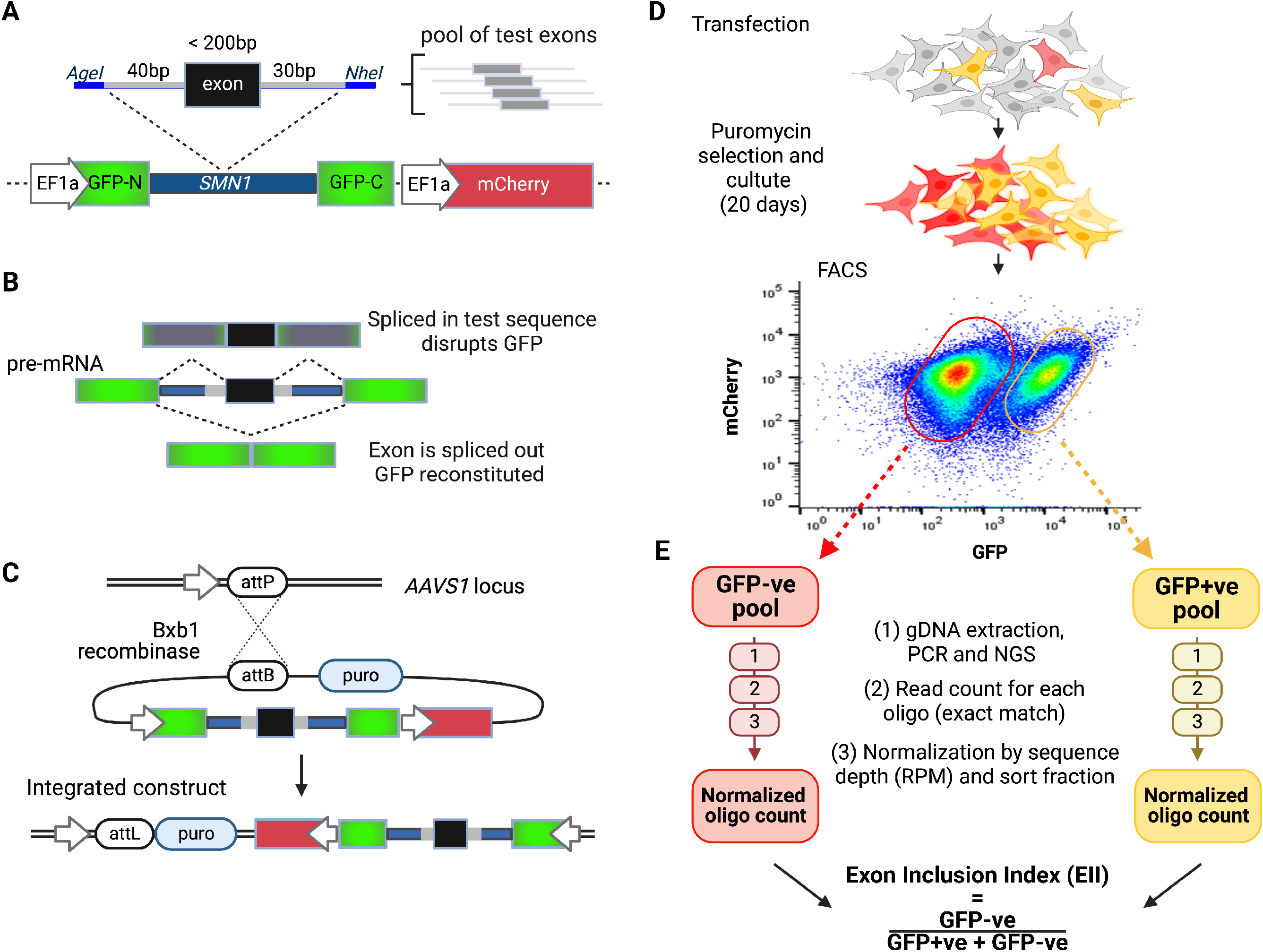
High throughput splicing assay (HTSA) experimental design and analysis. **A)** HTSA construct contains *GFP* split by an *SMN1* intron, into which a test exon is cloned by *AgeI* and *NheI* restriction cloning. mCherry is expressed independently as a background fluorescence control. A pool of exons (up to 200 bp in size) and flanking intronic sequences are cloned into a sequence derived from *SMN1* intron 7 and 8. **B)** Major splicing outcomes with exon inclusion leading to GFP disruption and exon exclusion leading to GFP reconstitution. Other outcomes are possible, e.g. partial exon inclusion or intron retention wich will also lead to the GFP disruption. **C)** RCA7 HEK293T landing pad locus contains an attP site and a gene-less promoter and the HTSA construct contains split GFP-SMN1 splicing minigene, mCherry and a promotor-less puromycin resistance gene. Co-transfection of the HTSA construct and a plasmid containing the Bxb1 integrase facilitates integration of one HTSA construct into the landing pad via the attB | attP pairing. Successful integration enables expression of puromycin resistance gene. **D)** RCA7 HEK293T landing pad cells are transfected with the HTSA plasmid pool and Bxb1 integrase. Selection with puromycin ensures elimination of cells without the integrated HTSA construct and prolonged cell growth (20 days) ensures elimination of episomal HTSA plasmids. Fluorescence activated cell sorting (FACS) leads to a clear separation of GFP+ve and GFP-ve cells. **E)** After isolation of gDNA from the cells, the integrated constructs are PCR-amplified and deep sequenced. The sequence reads are matched to the library sequences and read of each integrated sequence is counted in each cell population separately. The oligo counts are subsequently normalized by the sequence depth of the sample (reads per million, RPM) and by the FACS sorting cell fraction of the sample. The resulting normalized oligo cell counts are then used to calculate exon inclusion index as shown.

Other elements of the HTSA are analogous to MFASS^54^. The multiplexed libraries are integrated into the *AAVS1* safe harbor locus of the RCA7-HEK293T landing pad human cell line by co-transfection with a plasmid carrying the *Bxb1* integrase (Figure 1C)^54,84^. The integrated plasmid carries a puromycin resistance gene, which can only be expressed after integration of the HTSA construct in the *AAVS1* locus. Puromycin selection and multiple passages over 20 days of cell culture ensure that a single copy of the reporter construct is expressed per cell and episomic HTSA plasmids are degraded. This enables fluorescence activated cell sorting (FACS) separation of GFP negative (-ve) and GFP positive (+ve) cells (Figure 1D). The sequences in the GFP-ve and GFP+ve cell pools are subsequently analyzed by deep next generation sequencing, and the matching sequences counted and normalized by the read depth and by the fraction of cells sorted into the GFP+ve or GFP-ve bins. To determine which variants lead predominantly to exon inclusion or exclusion, the exon inclusion index (EII) is calculated for each sequence using the normalized oligo sequence count, where EII= GFP-ve read count/(GFP-ve + GFP+ve read count) (Figure 1E). EII of 1 signifies complete exon inclusion and EII of 0 indicates complete exon skipping.

### Evaluation of HTSA based on 380 IRD gene exons and essential splice site controls

In the initial experiment we tested 380 exons from 89 IRD genes that were chosen based on rare unclassified mutations found in unsolved IRD patient cohort from the MEE clinic (MAF < 0.003) and additional rare variants found in the gnomAD database (MAF < 0.001). The control library consisted of the reference exon sequences and two control sequences for each exon with mutated acceptor site (AS) or donor site (DS) (Supplementary Table 1). Owing to the improvement of the oligo synthesis methodology we were able to investigate exons up to 200 bp in length (median exon length 132 bp) compared to 100 bp long exons studied previously^54^.

Of the 380 exons, fifty were not sufficiently represented in both replicates (< 50 cells per sequence) and were dropped from the subsequent analyses. For the remaining 330 exons, we measured reproducibility of the assay across two biological replicates using the EII values. The data showed a bimodal distribution with EII values clustering at 0 - 0.1 (complete or high exon skipping) and 0.9 - 1 (complete exon inclusion), and a high level or reproducibility between the replicates (simple linear regression r^2^ = 0.83) (Figure 2A).

**Figure 2.**
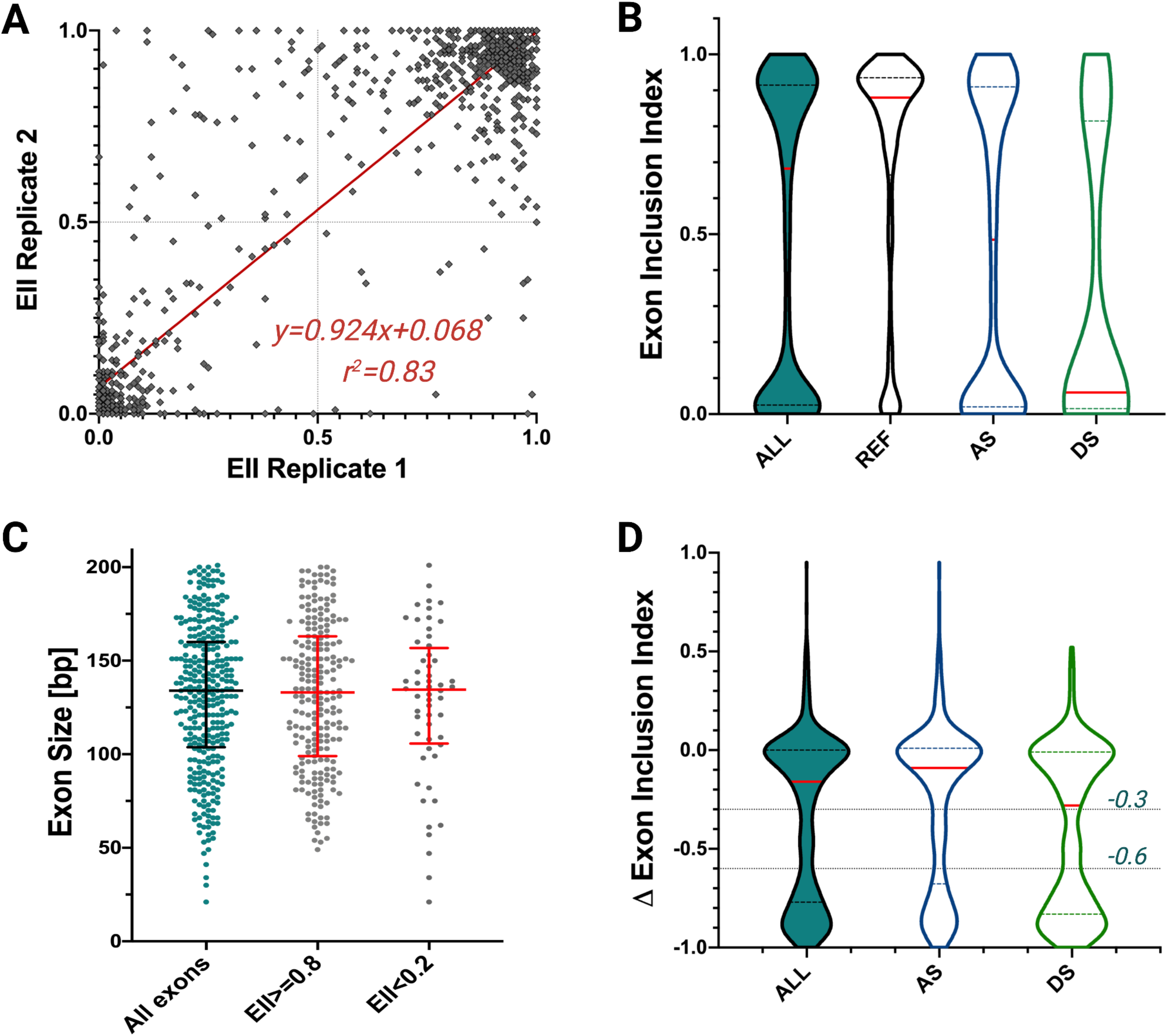
HTSA performance on a control library. **A)** Reproducibility of exon inclusion indexes (EII) from replicate 1 and 2 (linear regression correlation coefficient r^2^=0.83). **B)** Exon inclusion indexes for all control library sequences and separately for reference, acceptor (AS) and donor (DS) splice site mutations. **C)** Distribution of the overall exon lengths included in the study, median length = 132 base pairs (bp) and of the reference exon sequences that showed high rate of exon recognition (EII ≥ 0.8) or preferential exons skipping (EII < 0.2). **D)** ΔEII values for the essential splice site controls, demonstrating a bimodal distribution of variants around ΔEII = 0 and ΔEII = −1. The distribution of data suggests two cutoff point to estimate splicing effects: ΔEII≤ −0.3 for moderate effect on exon inclusion and ΔEII ≤ −0.6 for strong effect on exon inclusion.

Most of the reference exons showed a high level of exon recognition (median EII = 0.88) with 260 (79%) reference sequences recognized as exons at least half of the time (EII ≥ 0.5) (Figure 2B). Fifty-six (17%) reference sequences showed preferential exon skipping (EII<0.2) and 14 showed lower than expected exon recognition (0.2 ≤ EII < 0.5). There was no correlation between the length of the studied exons and their performance in the splicing assay (Fig. 2C, unpaired T-test: p-value = 0.5).

Introducing mutations to the essential splice sites reduced exon recognition, where DS controls showed stronger effect (median EII=0.06) than AS controls (median EII=0.48) (Figure 2B). To evaluate how essential splice site controls performed in comparison to their corresponding references, we used a ΔEII metric, where ΔEII = EII_control_ - EII_reference_. The ΔEII values clustered in two major groups: around ΔEII = −1 and around ΔEII = 0, where ΔEII = −1 indicates a complete exon skipping due to the mutation in a well performing reference exon, and ΔEII = 0 indicates no effect of variant on exon inclusion (Figure 2 D). In a few extreme cases ΔEII showed positive values, which indicated a stronger exon inclusion in sequences with an essential splice site variant than in the corresponding reference sequence (Figure 2 D). DS controls showed on average higher effect on exon recognition (median ΔEII = −0.28), compared to AS controls (median ΔEII = −0.09). Based on the bimodal distribution of the ΔEII values, we have determined that ΔEII ≤ −0.3 is a reliable cutoff for variants that show effect on exon inclusion and ΔEII ≤ −0.6 is a reliable cutoff for a strong effect on exon inclusion (Figure 3D).

**Figure 3.**
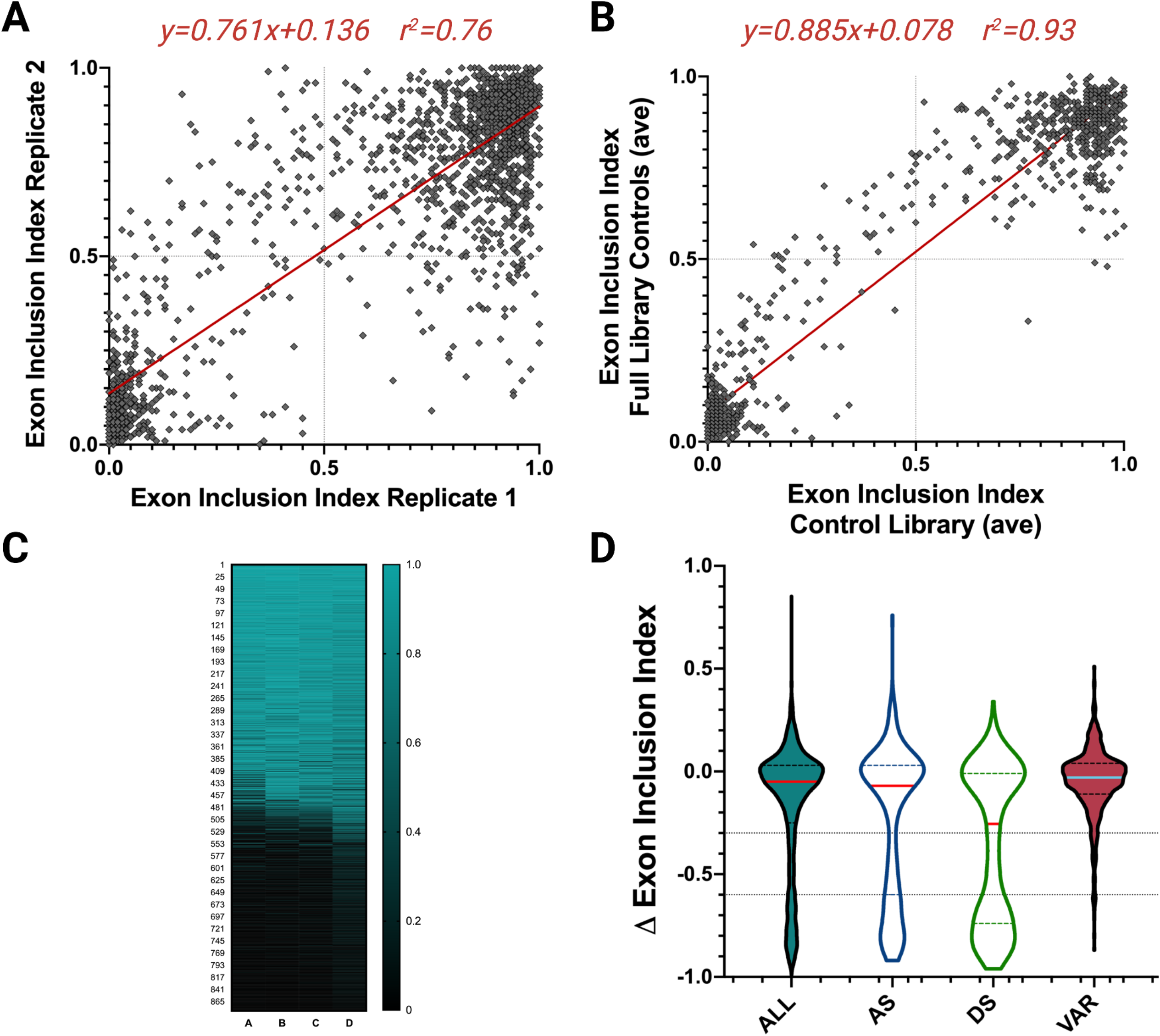
Measuring the effects of variants of unknown significance on exon recognition. **A)** Reproducibility of exon inclusion indexes (EII) from replicate 1 and 2 from a library containing 2166 different sequences of references, essential splice site controls and VUSs (linear regression correlation coefficient r^2^=0.76). **B)** A heatmap showing reproducibility of EII in control sequences (reference and essential splice site controls) from four biological experiments. **C)** Reproducibility of the averaged exon inclusion indexes (EII) for the control library and the control sequences from the full library (linear regression correlation coefficient r^2^=0.93) **D)** Delta exon inclusion indexes (ΔEII) for variant library sequences (essential splice site controls and tested variants) and separately for acceptor (AS), donor (DS) and tested variants (VAR).

### Evaluation of the effect on splicing of 1131 variants in IRD genes

Next, we tested variants of unknown significance in IRD genes. The variant library consisted of 380 exons, 785 essential splice site controls and 1131 test variants (2296 sequences in total) (Supplementary Table 2). First, we determined the reproducibility of the assay comparing two biological replicates. Since, our variant library contained over a thousand variants more than a control library, the correlation between the two replicates was lower for the same inclusion criteria (≥ 50 cells per sequence, r^2^ = 0.69), we therefore applied more stringent filtering and considered sequences that were present in at least 150 cells as determined by cell sorting and sequence coverage (Supplementary Table 2). With these criteria the two replicates were highly correlated (r^2^ = 0.76) (Figure 3A). Two hundred and ninety-six exons with 748 variants in the IRD genes passed this filtering threshold (Supplementary Table 2). Since the control library and the variant library contained the same reference and essential splice site control sequences, we were able to compare exon inclusion of both libraries across four experiments, demonstrating high reproducibility (Figure 3B). Pairwise comparison of the average values of EII for the control library and EII for the variant library were highly correlated, demonstrating robustness and reproducibility of the assay (r^2^ = 0.93, Figure 3C). As in the control library before, the DS controls exerted on average a higher impact on exon recognition (median ΔEII = −0.26) compared to AS controls (median ΔEII = −0.07). The overall effect of tested variants on exon inclusion was smaller than that of the positive controls (median ΔEII = −0.03) (Figure 3D).

Of the 748 tested variants, 47 changes (6.2%) reduced exon recognition by at least 30% (ΔEII ≤ −0.3) and eleven of these variants (1.4%) showed a strong effect on exon inclusion, reducing exon recognition by 60% or more (ΔEII ≤ −0.6) (Table 1, Supplementary Table 2). These included three synonymous variants, two missense changes and six intronic variants. Five of the variants showing a strong exon skipping effect were predicted to have a significant effect on splicing by the Splice AI algorithm (ΔSplice AI score >0.2)^55^ (Table 1).

**Table 1.** Variants showing a strong effect on exon inclusion.

| Gene name | Transcript ID | Exon | cDNA change | Protein change | gnomAD | Average $\Delta EI$ | SpliceAI | Exon frame |
| --- | --- | --- | --- | --- | --- | --- | --- | --- |
| *ABCA4* | ENST00000370225.3 | 47 | c.6438C>T | p.(=) | 0.000039 | -0.61 | 0.03 | In-frame |
| *CEP164* | ENST00000278935.3 | 10 | c.1233+5G>A | intronic variant | 0.000007 | -0.87 | **0.95** | In-frame |
| *CHM* | ENST00000357749.2 | 7 | c.940G>A | p.(Gly314Arg) | Absent | -0.63 | **0.84** | Out-of-frame |
| *CPLANE1* | ENST00000651892.2 | 27 | c.5520T>G | p.(=) | Absent | -0.62 | 0.06 | Out-of-frame |
| *CRX* | ENST00000221996.7 | 3 | c.158A>G | p.(Glu53Gly) | Absent | -0.80 | 0.00 | Out-of-frame |
| *EYS* | ENST00000370621.7 | 6 | c.1056+3A>C | intronic variant | Absent | -0.71 | **0.94** | Out-of-frame |
| *EYS* | ENST00000370621.7 | 6 | c.863-15T>C | intronic variant | 0.000004 | -0.64 | 0.00 | Out-of-frame |
| *EYS* | ENST00000370621.7 | 6 | c.863-5T>G | intronic variant | 0.000029 | -0.68 | 0.12 | Out-of-frame |
| *EYS* | ENST00000370621.7 | 6 | c.969A>G | p.(=) | 0.000007 | -0.61 | 0.00 | Out-of-frame |
| *EYS* | ENST00000370621.7 | 28 | c.5836-11T>G | intronic variant | 0.0000069 | -0.61 | **0.35** | Out-of-frame |
| *RPGR* | ENST00000645032.1 | 4 | c.310+7T>G | intronic variant | Absent | -0.65 | **0.37** | In-frame |

Notes: \*13 hemizygotes are present in gnomAD, indicating that partial defect in exon inclusion is not pathogenic or leads to a mild later onset disease, overlooked in the gnomAD cohort.

### New genetic solutions in IRD patients

When assessing pathogenicity of genetic variants that lead to exon skipping, we have to consider multiple criteria: 1) the frame of the exon that is eliminated; 2) protein domains that are encoded by the in-frame exon; 3) mode of inheritance of a particular trait and the pathogenicity of the allele in *trans* for the recessive genes; 4) gene dosage sensitivity^85^; 5) extent of exon skipping (full or partial). These criteria will allow us to determine if a particular variant leads to a hypomorphic or to a null allele. For example, partial skipping of an out-of-frame exon will lead to a hypomorphic allele, which can be pathogenic in a recessive gene, when coupled with a pathogenic or likely pathogenic variant in *trans* ^86,87^. Skipping of an in-frame exon, will not lead to transcript degradation but may lead to a non-functional protein if an important domain is deleted. Weighing these criteria, we evaluated variants that showed an effect on exon skipping in a cohort of ∼3000 IRD cases analyzed by targeted sequencing or exome sequencing^13,38^. We found five cases for whom the splicing results likely lead to new genetic diagnoses, including three cases with autosomal recessive gene solutions (*ABCA4*^88,89^, *EYS*^90,91^) and two with X-linked disease (*CHM*^94^, *RPGR*^95^) (Table 2).

**Table 2.** New likely genetic solutions.

| Patient number | Gene | gDNA Hg38 | HGVSc | HGVSp | gnomAD v3 | CADD | ClinVar | Average ΔElI |
| --- | --- | --- | --- | --- | --- | --- | --- | --- |
| **OGI3781_0052218** | *ABCA4* | chr1:94010821C>T | c.5693G>A | p.Arg1898His | 0.00147 | 19.52 | Conflicting interpretations | -0.45 |
|  |  | chr1:94031110G>A | c.4139C>T | p.Pro1380Leu | 0.0002431 | 25.3 | P/LP | N/A |
| **OGI2923_004508** | *ABCA4* | chr1:94111517A>G | c.223T>C | p.Cys75Arg | absent | 29.8 | LP | N/A |
|  |  | chr1:94010834G>A | c.5680C>T | p.(=) | absent | 5.195 | B* | -0.34 |
| **OGI3782_0052219** | *CHM* | chrX:85957855C>T | c.940G>A | p.(Gly314Arg) | Absent | 33 | VUS/LP** | -0.63 |
| **OGI2026_003452** | *EYS* | chr6:64230343-64231073:1 | Exon 31 heterozygous deletion |  | Absent (gnomADv2-SV) | N/A |  | N/A |
|  |  | chr6:65405171T>G | c.1056+3A>C |  | absent | 22.7 |  | -0.71 |
| **OGI1347_002520 (male)** | *RPGR* | chrX:38321020A>C | c.310+7T>G |  | absent | 10.65 | VUS | -0.62 |

\*Benign by one submitter; \*\* classification if this variant has recently been changed from VUS to LP in CLinVar. \*\*\*this variant is not thought to act via exon skipping but it is predicted to be likely damaging by PolyPhen, fathmm and has an elevated CADD and Revel (0.5) scores

Two subjects carried mutations in *ABCA4*, a gene associated with a number of retinal disorders including an inherited macular degeneration (OMIM #248200), cone-rod (OMIM #604116) and rod-cone dystrophy (OMIM #601718)^88,89^. The studied cases were affected with cone-rod dystrophy (OGI3781_0052218) and macular degeneration (OGI2923_004508) and they had heterozygous variants leading to an out-of-frame exon 40 skipping, which is thought to result in nonsense-mediated decay^46^. Variants c.5693G>A, p.Arg1898His (ΔEII = −0.45) and c.5680C>T, p.Leu1894= variant (ΔEII = −0.33), both lead to a partial exon 40 skipping. These alleles can be considered as hypomorphic alleles, which have been shown to lead to *ABCA4*-associated disease^86^. In both cases the hypomorphic splicing variants were coupled with pathogenic/likely pathogenic variants c.4139C>T, p.Pro1380Leu and c.223T>C, p.Cys75Arg respectively (Table 2)^98,100,101^.

A male subject OGI3782_0052219 with a diagnosis of an X-linked condition of choroideremia carried a hemizygous variant c.940G>A, p.(Gly314Arg) in *CHM*, a gene associated with the subject’s condition (OMIM #303100)^94^. The c.940G>A variant leads to skipping of an out-of-frame exon 7 in 2/3^rd^ of the transcripts (ΔEII = −0.63), is absent in gnomAD and segregated with the same phenotype in a maternal male cousin and thus is likely to lead to disease. At the time of the study the variant was classified as VUS, however recently the classification has changed to likely pathogenic, which corroborates our findings^98^.

A rod-cone dystrophy case OGI2026_003452 carried two variants in *EYS*, associated with a recessive form of IRD (OMIM # 602772) ^90,91^. The subject carried a heterozygous deletion of an out-of-frame exon 31 and a c.1056+3A>C change leading to skipping of an out-of-frame exon 6 in over 2/3^rd^ of the transcripts (ΔEII = −0.71). These two changes are a very likely cause of disease in this subject (Table 2).

A simplex male patient with rod-cone degeneration was found to carry a hemizygous c.310+7T>G variant in *RPGR*, a gene associated with an X-linked IRD (OMIM # 312610)^95^. The c.310+7T>G variant affected splicing of an in-frame exon 4, leading to exon skipping in up to 2/3^rd^ of transcripts (ΔEII = −0.62). Exon 4 codes for 21 residues that are involved in forming of the second regulator of chromosome condensation (RCC1) domain^102,103^ (Table 2).

## DISCUSSION

In this study we have developed an improved version of a high-throughput splicing assay (HTSA), which was based on a previously published method^54^. We used HTSA to study 1131 variants of unknown significance in 380 exons from 89 IRD genes. However, due to a limited library diversity, we were able to study only 748 variants. We observed that 47 (6.2%) variants lead to decreased exon inclusion. Of these, 11 variants showed a strong effect, with at least 60% of transcripts leading to exon skipping compared to the reference sequences. Using the splicing assay data we were able to provide a likely genetic diagnosis to ten IRD cases.

HTSA showed high reproducibility when compared between different biological replicates. Most of the reference exons showed a high level of exon recognition (median EII = 0.88), however in 70 reference sequences the exon was recognized less than 50% of the times and 56 exons were preferentially skipped (EII<0.2). One of the reasons for the poor performance of these exons could have been the lack of the crucial intronic context, necessary for the exon recognition. However we have not observed a correlation between the size of a studied exon (the smaller the exon the larger the flanking exonic sequences in the 270bp oligos) and its performance in the splicing assay.

We have detected 11 variants with strong effects on splicing, however not all of these variants led to genetic solutions in our patient cohort, which can be due to a number of reasons. In some autosomal recessive cases the second variant in the same gene was determined not to be pathogenic, a phenotype didn’t match or a another more likely cause of disease was found. Nevertheless in three recessive cases, our HTSA was able to provide additional functional evidence to reconsider variants of unknown significance that were coupled with likely pathogenic variants in the same gene. In two X-linked male cases, exon skipping led to the deletion of a crucial domain (RCC1 in *RPGR*) or creation of a frameshift mutation (*CHM*), which were likely causal variants.

In HTSA we measured the disruption of GFP, which we labeled as exon recognition. However, disruption of GFP may also occur when part of an exon is included, or an intron retained, which can only be identified by the analysis of HTSA minigene transcript from the GFP-ve pool of cells. Therefore it is possible that variants that seemingly showed no effect on exon inclusion, actually led to aberrant splicing by one of the above mechanisms. Further studies will be undertaken to test this hypothesis. Of the 11 variants that showed a strong effect on exon inclusion, only five were predicted to have a significant effect on splicing by the Splice AI algorithm (ΔSplice AI score >0.2)^55^. This illustrates the need for improved splicing prediction programs especially in regions not typically thought of as splice altering. American College of Medical Genetics (ACMG) guidelines often require functional validation to assess the pathogenicity of unclassified variants^65^. However, the cost and effort to individually interrogate VUSs is often prohibitive which leaves many variants that may provide the genetic solution as unclassified. We present an elegant and robust method for interrogating hundreds to thousands of variants and their impact on exon recognition leading to splicing dysfunction. In the current study HTSA was employed to investigates the effects of variants in or near exons, however it can also be used to study deep intronic variants leading to cryptic exon inclusion^48–52,77^.

## Data Availability

The raw data from the splicing assay are provided in the supplementary files

## ACKNOWLEDGMENTS

This work was supported by grants from the National Eye Institute R01EY026904 (KMB/EAP), [R01EY012910 (EAP) and P30EY014104 (MEEI core support)], the Foundation Fighting Blindness [EGI-GE-1218-0753-UCSD, (KMB)] and the Research to Prevent Blindness International Research Collaborators Award (KMB). Exome sequencing and analysis were provided by the Broad Institute of MIT and Harvard Center for Mendelian Genomics (Broad CMG) and was funded by the National Human Genome Research Institute, the National Eye Institute, and the National Heart, Lung and Blood Institute grant UM1HG008900 and in part by National Human Genome Research Institute grant R01 HG009141. The authors thank all subjects for their participation in this study and the OGI Genomics Core members for their experimental assistance.

